# HOW COMPLETE ARE TOBACCO SALES DATA? ASSESSING THE COMPREHENSIVENESS OF TOBACCO PRODUCT RETAIL SALES DATA THROUGH COMPARISONS TO EXCISE TAX COLLECTIONS

**DOI:** 10.1101/2023.05.01.23289351

**Authors:** Alex C. Liber, Maryam Faraji, Radhika Ranganathan, Abigail S. Friedman

## Abstract

**Introduction:** Sales data analyses are increasingly used to guide tobacco regulatory science. However, such data do not cover specialist retailers like vape shops or tobacconists. Understanding the extent of the cigarette and electronic nicotine delivery system (ENDS) markets covered by sales data is critical to establishing such analyses’ generalizability and potential biases.

**Methods:** Sales data from Information Resources Incorporated (IRI) and Nielsen Retail Scanner data are used to conduct a tax gap analysis, comparing state tax collections based on cigarette and ENDS sales data to states’ annual 2018-2020 cigarette tax collections and monthly ENDS and cigarette tax revenue data for January 2018 to October 2021. Cigarette analyses consider the 23 US states covered by both IRI and Nielsen. ENDS analyses consider the subset of those states with per unit ENDS taxes: Louisiana, North Carolina, Ohio, and Washington.

**Results:** Across states covered by both sales datasets, IRI’s mean cigarette sales coverage was 92.3% (95% CI 88.3-96.2%), while Nielsen’s was 84.0% (95% CI 79.3-88.7%). Coverage rates for average ENDS sales were lower, ranging from 42.3% to 86.1% for IRI and 43.6% to 88.5% for Nielsen, but remained stable over time.

**Conclusions:** IRI and Nielsen sales data capture almost the entire US cigarette market and, while coverage rates are lower, a substantial portion of the US market for ENDS as well. Coverage rates are relatively stable over time. Thus, with proper care to address shortcomings, sales data analyses can capture changes in the US market for these tobacco products.

**Implications:**

- Policy evaluations and analyses using e-cigarette and cigarette sales data are often criticized because these data do not cover online sales or sales by specialty retailers like tobacconists.
- Cigarette sales data consistently cover nearly 90% of taxed sales, while e-cigarette sales data cover around 50% of taxed volumes.
- Retail sales data capture nearly all cigarette sales and a substantial portion of ENDS sales with relatively stable rates of coverage over time, supporting their continued use in tobacco surveillance and policy evaluation work.

## Introduction

Researchers are increasingly using retail sales data sold by companies like Information Resources Incorporated (IRI) and Nielsen IQ to assess tobacco policies’ effects and inform regulation: over 90 such manuscripts have been published to date^A^. This data is passively compiled over time from barcode scanners at mainstream brick-and-mortar. These sources yield rapidly accessible, geocoded data and enable timely and precise tobacco market surveillance and policy evaluation research.

Concurrently, sales data are often criticized for presenting an incomplete view of tobacco markets. They do not cover online retailers, stores on Native American reservations, or specialty stores such as tobacconists and vape shops. Moreover, the relative importance of uncovered retailers varies across geographies due to differences in licensing regulations and the presence of reservations within a state.^1^

Consequently, using retail sales data as a tobacco surveillance and policy evaluation tool requires a clear understanding of its representativeness, yet few studies have assessed this. Purveyors of sales data estimate that they cover approximately 77% of all retail stores in the US.^2^ However, market research firms estimate that more than half of US electronic nicotine delivery system (ENDS) sales occur in retail channels missing from these datasets.^3^ Financial analysts have estimated the value share of US tobacco product sales from online stores ranges from 15% to 35%, with growth over the past decade,^3^ and that a similar or smaller share of ENDS sales occurs in vape shops, with that share declining in recent years.^4^ However, the methods behind these estimates are opaque and subject to retroactive revision.

To interrogate retail sales data’s potential as a surveillance and evaluation instrument for the tobacco products marketplace, this research presents modified tax gap analyses that estimate the percent of cigarette and ENDS consumption captured by IRI and Nielsen, the two leading sources of US retail scanner data on these products. Specifically, we compare state cigarette and ENDS sales data recorded by IRI and Nielsen to total sales implied by state tax rates and excise tax receipts, considered the most thorough accounting of state tobacco market size.

Consideration is limited to states with data from both sources and, for ENDS, states with per unit ENDS taxes, where ENDS sales volume in milliliters can be deduced from tax receipts. Results provide a concrete comparison to establish the true extent of sales data’s cigarette and ENDS market coverage. Due to the relative importance of vape shops and online retailers in the ENDS marketplace, we expect retail sales data on ENDS to cover a lower proportion of tax collections than for cigarettes, which rely almost exclusively on sales in mainstream retailers.

## Methods

We obtained retail sales data from Information Resources Inc (IRI) for January 2018 - March 2023 and Nielsen Scantrack for January 2017 - November 2021. Sales in all major brick-and-mortar retailers were present in both datasets from 23 states (see Figure 1). Annual state cigarette tax collections came from the *Tax Burden on Tobacco* (TBOT).^5^

**Figure 1:**
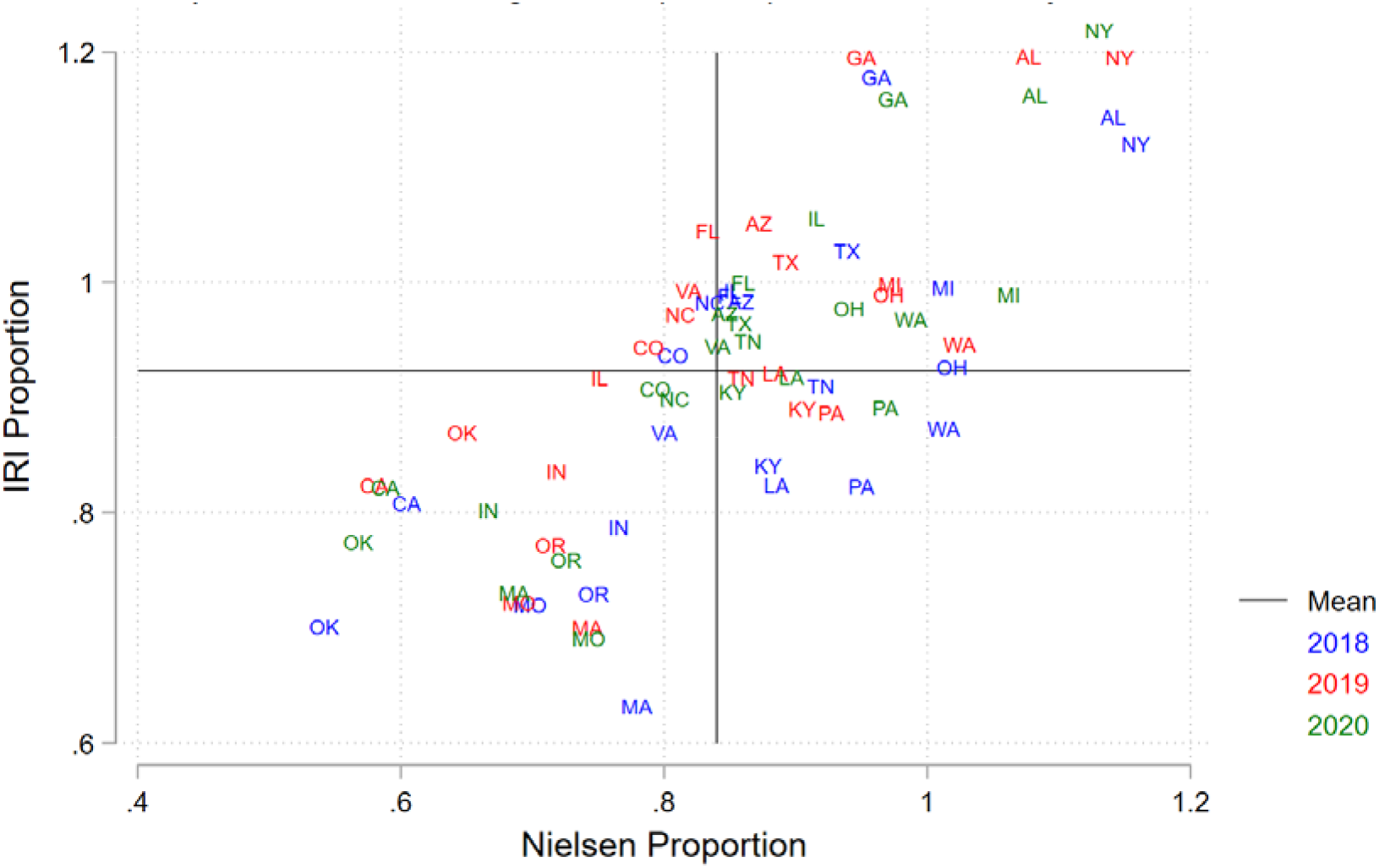
Proportion of Taxed Cigarettes in TBOT in Sales Data by Source, State, and Year

ENDS tax collection analyses considered the subset of those 23 states that imposed only specific ($/mL) taxes prior to September 2022: Louisiana ($0.05/mL since July 1, 2015), North Carolina ($0.05/mL since June 1, 2015), Ohio ($0.10/mL since October 19, 2017), and Washington ($0.27/mL for ENDS with less than 5 milliliters of e-liquid and a $0.09/mL for those with more than 5 milliliters of e-liquid, as of October 1, 2019).^6^ We submitted Freedom of Information Act (FOIA) requests to Louisiana, North Carolina, Ohio, and Washington for monthly excise tax collections of all tobacco products, disaggregated by states’ product categories, through October 2021. All complied except Ohio, which provided monthly ENDS tax collection data up through July 2021 and monthly cigarette tax collection data through October 2021, but an annual lump sum for fiscal year 2022 collections. After dividing Ohio’s 2022 collection into 12 equal portions to serve as a denominator from July 2021 to October 2021, we had complete ENDS tax and sales data coverage for January 2018 to October 2021.

We perform a modified tax gap analysis. Often used to estimate illicit cigarette consumption,^7,8^ this technique typically estimates total cigarettes consumed by multiplying average cigarette consumption per person by the population cigarette smoking prevalence and then subtracting off tax-collection-imputed legal sales to obtain the size of the illicit market. Instead of subtracting survey-based consumption estimates, we use the aforementioned IRI and Nielsen scanner-measured sales data to clarify the percentage of taxed cigarette and ENDS sales captured by those datasets.

Specifically, sales data information on sales volume, product size, and date of sale was combined with state tax rates to deduce the total tax revenue attributable to IRI and Nielsen cigarette and ENDS sales by state and calendar month. Dividing those figures by excise tax collection figures yielded the proportion of states’ cigarette and ENDS tax collections accounted for by sales documented in IRI and Nielsen data. We then used linear panel regressions with month and state-fixed effects to assess whether this coverage or inflation-adjusted tax collections changed over time. Additional models were run to determine the effect of expanding the March 2021 PACT Act amendments that banned the shipping of ENDS through the US Postal Service, which may have reduced consumers’ reliance on online sales and increased state tax collections^9^.

## Results

Figure 1 plots the share of 2018, 2019, and 2020 cigarette tax revenue explained by IRI data against the share explained by Nielsen data for each state covered in both datasets. Variation in coverage rates is evident across states more so than data sources. For the 23 states covered in both IRI and Nielsen with both all channel and convenience store data, the mean coverage of cigarette sales is 92.3% (95% CI 88.3-96.2%) and 84.0% (95% CI 79.3-88.7%) for IRI and Nielsen, respectively. The largest difference between IRI and Nielsen’s coverage rates for a given state is 24.5 percentage points (Mean absolute difference 11.6%, 8.4-14.5% 95% CI). Concerningly, sales data in Alabama and New York explain more than 100% of their excise tax collections in both years and datasets, potentially reflecting modeling decisions by sales data vendors.

Focusing on states with specific taxes on ENDS, Figure 2 shows the tax revenue generated by cigarette and ENDS sales in the IRI and Nielsen data over time as a percentage of actual state tax collections reported in FOIA requests. While cigarette sales coverage rates hovered around 95%, average ENDS coverage rates for IRI and Nielsen, respectively, were 55.3% and 44.8% in LA, 42.3% and 43.6% in NC, 86.1% and 88.5% in OH, and 62.8% and 67.5% in WA. Pooling across all four states, the model does not find significant changes in coverage of ENDS or cigarette sales over time but does indicate time-invariant differences (Supplemental Tables 1 and 2). Adding a control for the PACT Act shows that this policy change was associated with significantly higher ENDS tax collections alongside significantly lower ENDS tax coverage in both the sales data sources.

**Figure 2:**
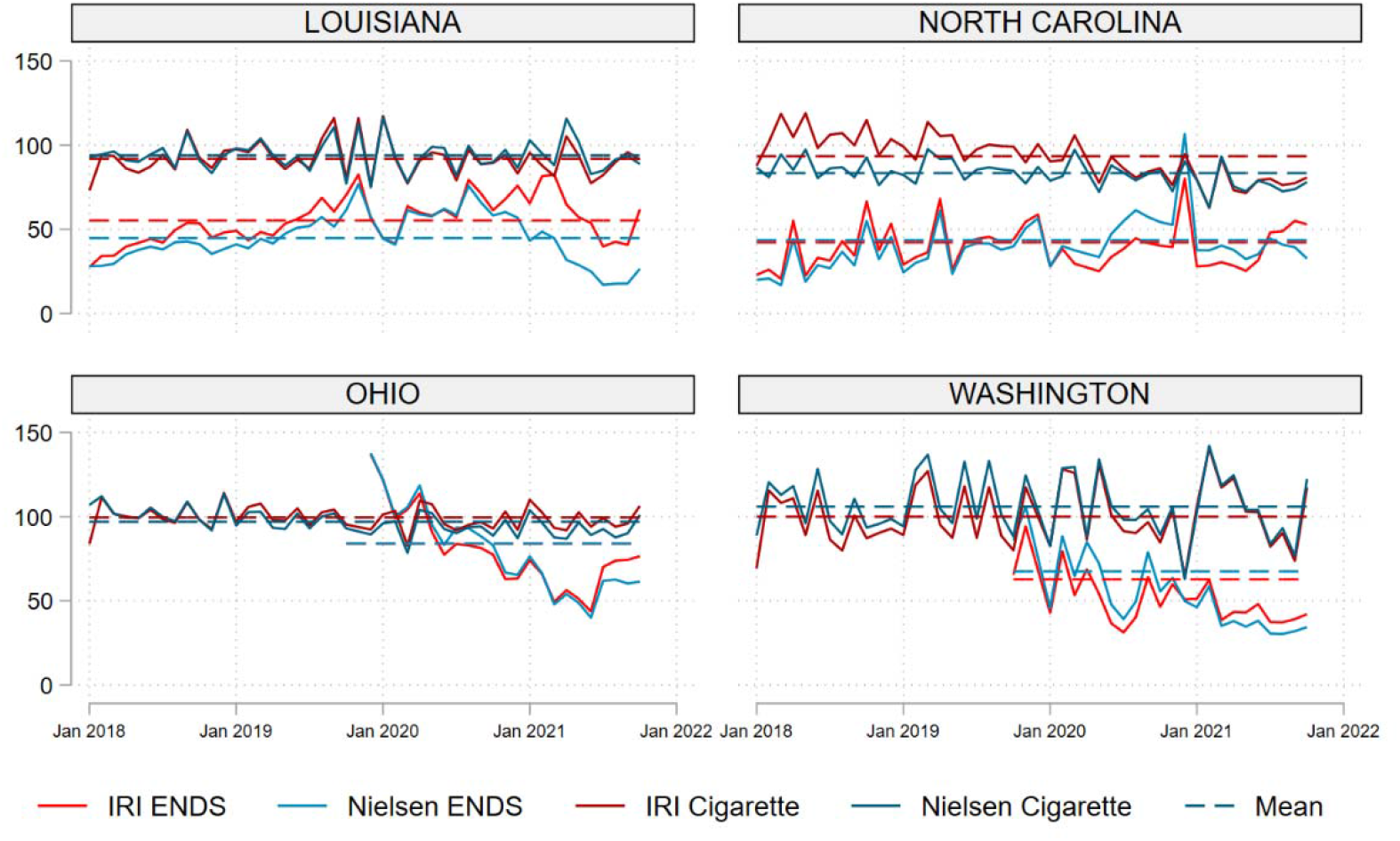
Volumes Sales as a Proportion of Monthly ENDS and Cigarette Excise Tax Collections from FOIA Requests **Notes:** IRI e-cigarette tax collections were higher than 100% of actual collections in 0 of 46 observations in LA, 1 of 46 observations in NC, 5 of 20 observations in OH, and 1 of 25 in WA. Nielsen e-cigarette tax collections were higher than 100% of collections in 0 of 46 observations in LA, 2 of 46 observations in NC, 6 of 20 observations in OH, and 2 of 25 in WA. IRI cigarette tax collections were higher than 100% of actual collections in 7 of 46 observations in LA, 15 of 46 in NC, 21 of 46 in OH, and 22 of 46 in WA. Nielsen cigarette tax collections were higher than 100% of actual collections in 8 of 46 observations in LA, 1 of 46 in NC, 16 of 46 in OH, and 25 of 46 in WA.

## Discussion

To our knowledge, this is the first study to quantify the comprehensiveness of US ENDS data coverage in the leading sources of retail sales data relative to state tax collection data. As expected, ENDS market coverage is inferior to cigarette market coverage, with the latter nearer to 100%. Both excise tax collection sources (TBOT and FOIA) yield similar estimates. Reassuringly, the overall coverage of ENDS sales is high in both datasets and does not appear to be changing over time. Moreover, the share of ENDS sales represented by monetary value may be even higher than estimates by volume since the per-unit cost of a milliliter of e-liquid purchased from mainstream retailers is much higher than that of e-liquid purchased in a vape shop.^10^ The introduction of the PACT Act, however, was associated with increases in ENDS tax collections alongside decreases in retail sales data coverage of that market. The increase in tax revenue without concomitant increases in retail sales data might be explained if some consumers who previously purchased ENDS products on the internet substituted towards brick-and-mortar retailers not covered by IRI and Nielsen, such as vape shops and tobacconists.

These findings suggest that retail sales data can be used to assess cigarette and ENDS sales changes in response to specific policies, as long as the policies in question do not substantively impact where people purchase these products. The analysis adjusts for time-invariant differences between states. Further work is needed to understand policies’ effects on sourcing from mainstream brick-and-mortar retailers versus online and specialty stores, particularly for ENDS.

### Limitations

Questions about data limitations and generalizability limit the conclusions of this analysis. First, monthly state excise tax collections do not perfectly align with our time periods for retail sales data; the latter is provided for four-week periods. We address this by splitting four-week sales period data to match calendar months, assuming sales on each day of a given period are equal. That simplification may add noise but should not bias overall estimates. Second, we do not cover cigars as only three states in the IRI and Nielsen data have per-unit cigar taxes, and applied tax rates vary across cigar types in a manner that is not reliably assessable from package attributes.^11^ This prevents us from deducing differences in sales volumes from excise tax revenue. Third, findings that sales data account for over 100% of cigarette tax revenue in some states raise questions about retail sales sources. These discrepancies could reflect vendors’ modeling decisions designed to capture sales in outlets where they do not collect data.^12^ Further work is needed to explain this. Fourth, we do not adjust for the effects of policy changes beyond the PACT Act amendments, consequently excluding the 2019 outbreak of vaping-associated lung injuries and the COVID-19 pandemic. These may impact the retailer dataset’s coverage of total sales if they affect where people purchase tobacco products (e.g., increased reliance on online sales during COVID lockdowns). More work is needed to clarify how such policy changes affect ENDS sourcing and potential implications for retail sales data.

Finally, online stores may not pay ENDS taxes in the states where their products are consumed, potentially creating a gap between tax-paid ENDS sales and the total market. The Supreme Court’s June 2018 decision in South Dakota v Wayfair—finding that states can mandate online sales tax collections from retailers that do not have a physical presence in that state—may alleviate this concern in more recent years.^13^ Future work quantifying the size of state ENDS markets to account for online sales and assessing how these vary over time would be valuable.

## Conclusions

Evidence that retail sales data cover a substantial portion of ENDS and nearly all cigarette sales supports the continued use of such data in tobacco surveillance and policy evaluation work. Analysts should adjust for time-invariant differences between states and account for potential effects on product sourcing. Further work is needed to assess policies’ effects on product sources at both the national and state level, to inform analyses that use retail sales data, and clarify policies’ implications for public health.

## Data Availability

The authors are not permitted to share the data obtained from NielsenIQ or Information Resources Inc with third parties.

## Footnotes

A PubMed search on 3/28/2023 using term: ((“tobacco*”[ Title/Abstract] OR “Cigarette*”[ Title/Abstract] OR “Cigar*”[ Title/Abstract] OR “e-cigarette*”[ Title/Abstract]) AND (ACNielsen*[Title/Abstract] OR Nielsen*[Title/Abstract] OR “Information Resources, Inc”[Title/Abstract] OR “retail sales”[Title/Abstract] OR “scanner data”[Title/Abstract]) NOT gene* found 126 manuscripts, of which 91 used the relevant data. The authors found additional manuscripts outside of PubMed and outside the above search terms.

