## Supplementary material for "HOW COMPLETE ARE TOBACCO SALES DATA? ASSESSING THE COMPREHENSIVENESS OF TOBACCO PRODUCT RETAIL SALES DATA THROUGH COMPARISONS TO EXCISE TAX COLLECTIONS": (Supplemental Tables 1 and 2)

### Supplementary Tables

Supplemental Table 1: ENDS Tax Collections and Market Coverage by Sales Data Source Regressed Against a Time Trend and The Introduction of the PACT Act Amendments

| **ENDS** | **Nielsen Coverage** | | **IRI Coverage** | | **Tax Collections** | |
| --- | --- | --- | --- | --- | --- | --- |
|  | **Time** | **+PACT** | **Time** | **+PACT** | **Time** | **+PACT** |
| **Time** | -0.00285 | 0.00426 | -0.000892 | 0.00247 | 29.73 | 12.75 |
| **PACT Act** |  | -0.363*** |  | -0.171* |  | 859.8*** |
| **State (LA Base)** |  |  |  |  |  |  |
| **NC** | -0.0117*** | -0.0117*** | -0.130*** | -0.130*** | 1121.0*** | 1121.0*** |
| **OH** | 0.411*** | 0.386*** | 0.286*** | 0.274*** | 1533.4*** | 1596.8*** |
| **WA** | 0.246*** | 0.219*** | 0.0734 | 0.0610 | 3040.9*** | 3107.0*** |
| **Mean** | 0.551 | 0.551 | 0.573 | 0.573 | 1947.47 | 1947.47 |

Note: p<0.001=***; p<0.01=**; p<0.05=*; The only covariates not listed above are month-fixed effects, included to adjust for seasonality. Standard errors are clustered by state.

Supplemental Table 2: Cigarette Tax Collections and Market Coverage by Sales Data Source Regressed Against a Time Trend and The Introduction of the PACT Act Amendments

| **Cigarettes** | **Nielsen Coverage** | | **IRI Coverage** | | **Tax Collections** | |
| --- | --- | --- | --- | --- | --- | --- |
|  | **Time** | **+PACT** | **Time** | **+PACT** | **Time** | **+PACT** |
| **Time** | -0.00145 | -0.00110 | -0.00169 | -0.00130 | -512.9** | -405.9** |
| **PACT Act** |  | -0.0213 |  | -0.0237* |  | -6601.4 |
| **State (LA Base)** |  |  |  |  |  |  |
| **NC** | -0.105*** | -0.105*** | 0.0153*** | 0.0153*** | -3213.2*** | -3213.2*** |
| **OH** | 0.0302*** | 0.0302*** | 0.0766*** | 0.0766*** | 193203.7*** | 193203.7*** |
| **WA** | 0.121*** | 0.121*** | 0.0811*** | 0.0811*** | 25927.2*** | 25927.2*** |
| **Mean** | .951 | .951 | .962 | .962 | 135185.1 | 135185.1 |

Note: p<0.001=***; p<0.01=**; p<0.05=*; The only covariates not listed above are month-fixed effects, included to adjust for seasonality. Standard errors are clustered by state.
